# Psilocybin and Contrasting Musical Compositions: Effects on Music-Evoked Emotional Responses in Healthy Adults

**DOI:** 10.64898/2026.09.09.26362648

**Authors:** Dominika Dojčánová, Filip Tylš, Marek Nikolič, Tomáš Novák, Vojtěch Viktorin, Anna Bravermanová, Michaela Viktorinová, Renáta Androvičová, Veronika Andrashko, Jakub Korčák, Peter Zach, Jiří Horáček, Martin Brunovský, Tomáš Páleníček

## Abstract

Music is a central component of psychedelic settings, yet little experimental work has examined how psilocybin and musical composition influence music-evoked emotional responses in healthy volunteers. This exploratory, double-blind, placebo-controlled study assessed these effects during the plateau phase of psilocybin and the corresponding time window in placebo sessions following oral administration of psilocybin (0.26 mg/kg) or placebo. The study consisted of two parts. In Part 1 (n = 19), a pilot stage, the first half of participants rated music-induced affective valence for eight compositions representing different genres/subgenres (Rock/Punk, Metal/Industrial Metal, Metal/Industrial Metal, Classical/Baroque, Electronic/Dub, Folk/Nordic Folk, Electronic/Techno, Electronic/Trance, Electronic/Psytrance) using a visual analogue scale (VAS). Based on Part 1, the classical composition with the highest overall affective valence and the psytrance composition with the largest psilocybin–placebo contrast (i.e., the composition most responsive to psilocybin) were selected for Part 2 (n = 17), in which the second half of participants assessed emotional responses to music using the Geneva Emotional Music Scale (GEMS-9). Psilocybin increased wonder, transcendence, and joyful activation across both contrasting compositions. Relative to each other, the classical composition elicited greater peacefulness, tenderness, and nostalgia, whereas the psytrance composition elicited greater tension across both psilocybin and placebo conditions. A composition × condition interaction was observed for sadness: under psilocybin, sadness increased during the classical composition and decreased during the psytrance composition. These findings suggest that psilocybin and contrasting musical compositions independently influence different aspects of music-evoked emotional responses, with limited interaction effects, and point to potential implications for protocol design and music selection in psychedelic settings.

## Introduction

Psilocybin, a classic serotonergic psychedelic (5-HT A receptor agonist), is a psychoactive compound found in “magic mushrooms,” with a long history of ritual use among Mesoamerican Indigenous communities [1,2]. In recent years, it has received growing attention in neuroscience and psychiatry for its potential to treat a range of neuropsychiatric conditions, particularly when combined with psychological support [3–8]. While psilocybin itself produces profound psychoactive effects, the overall psychedelic experience is also significantly shaped by “Set” and “Setting“ [9–11]. “Set” encompasses both long-term characteristics, such as personality traits, inner conflicts, and ambitions, as well as momentary states like current mood, expectations, and attitude, whereas “Setting” refers to the physical, social, and cultural context [12,13]. Among elements of “Setting“, music is especially prominent in psychedelic sessions, yet the precise ways in which it interacts with psychedelics to shape the experience remain only partly understood.

Two of the earlier modern studies on psychedelics using LSD revealed that it increased the overall emotional response to music and amplified music-evoked emotions such as wonder, transcendence, power, and tenderness [14], as well as enhanced music-induced visual imagery in healthy volunteers [15]. Few studies have explored music under psilocybin; to our knowledge, all conducted with depressed patients. Qualitative research indicates that music facilitates a shift from emotional avoidance to acceptance [16]. Results demonstrated both welcome and unwelcome effects of music on subjective experience, and perceptions of music, particularly liking, resonance, and openness, were associated with mystical-type experiences and predicted lower depression one week later, whereas drug-intensity ratings did not [17]. Music increased subjective pleasure independently of psilocybin, with no main effect of treatment or music × treatment interaction observed for pleasure ratings. At the same time, music-evoked emotional responses shifted following treatment, with increased peacefulness and reduced sadness. [18]. Extending these findings, in both the psilocybin and the active comparator (escitalopram) groups, stronger music-evoked emotion during sessions was associated with greater symptom improvement [19]. Overall, these findings indicate that music plays a central and clinically relevant role in shaping affective experience in psychedelic contexts, with music-related effects shown to operate also independently of pharmacological state. Evidence also suggests that psychedelics may alter sensitivity to emotionally salient stimuli such as music, yet clear interaction effects on affective responses have not been consistently observed to date.

Looking more closely at music, different genres shape distinct emotional profiles [20], suggesting that genre choice may be a practical means to modulate affect under psychedelics. Across contexts, communities that use psychedelics show characteristic musical preferences. In Indigenous settings, elements such as ícaros - healing songs [21] and drumming [22] are central to shamanic ceremonies. In Western culture, psilocybin is commonly associated with rave culture, characterized by electronic music genres such as techno [23] and psytrance [24,25]. Psilocybin was also linked with the hippie movement [26], which was closely associated with rock music styles, notably psychedelic rock [27]. Furthermore, playlists developed for psilocybin therapy often consist of a combination of genres, predominantly classical music [28], such as Western classical [29] and neo-classical, as well as ambient music [17]. This diversity indicates that musical genre is treated as an intentional and potentially functional component of psychedelic settings; however, whether and how different genres modulate affective experience under psilocybin remains incompletely characterized.

Patient reports suggest genre and musical features matter: tribal drum beats evoked vivid imagery, whereas darker tracks surfaced difficult emotions and sometimes overwhelming experiences [16]. Design guidance favors structured phrasing, consistent instrumentation, and a sense of forward motion to support peak experiences [30]. However, direct experimental comparisons of musical genres under psychedelics are rare. Notably, the only head-to-head comparison identified found no clear advantage on subjective or clinical outcomes between Western classical music and overtone-based music, including Tibetan singing bowls, gongs, didgeridoo, chimes, bells, sitar, and human voice overtone singing, during psilocybin-assisted smoking cessation therapy [31].

The main objectives of this exploratory study were to examine the effects of psilocybin and musical compositions from different genres/subgenres on music-induced affective valence and emotional responses in healthy volunteers. Specifically, we tested (i) the effect of psilocybin across compositions, (ii) the effect of composition (representing different genres) across psilocybin and placebo conditions, and (iii) their interaction. This focus is novel, as prior research on psilocybin and music has focused almost exclusively on clinical populations with little systematic attention to musical genre, limiting inferences about music-related effects that are independent of clinical status. These findings generate hypotheses about the role of musical context in psychedelic settings and offer preliminary insights into how psilocybin influences music-evoked emotional processing.

## Experimental procedures

### Approvals

The present study used data from a larger placebo-controlled trial evaluating behavioral and EEG/fMRI changes induced by psilocybin. The study was conducted at the National Institute of Mental Health (formerly the Prague Psychiatric Center). Regulatory authorization was granted by the State Institute for Drug Control on 11 December 2013 (registration no. 92/13), and the study was registered as a clinical trial under EudraCT No. 2012-004579-37. Ethics approval was obtained on 18 June 2014 (ref. 60/14) from the Internal Review Board (IRB), then titled the “Ethical Committee of the Prague Psychiatric Centre,” which later became the “Ethical Committee of the National Institute of Mental Health” and oversaw the study for its remainder. Participants were recruited from 1 October 2014, with the final participant visit completed on 3 October 2020. The study was conducted in accordance with the revised Declaration of Helsinki (2000) and the Guidelines for Safety in Human Hallucinogen Research (Johnson et al., 2008). All participants were fully informed and provided written informed consent (“Information for Volunteers and Informed Consent to Participate in a Clinical Research Study”).

### Study design

For a full description of the original trial design and patient selection, please refer to an earlier publication [32]. In brief, potential participants were initially pre-screened by a telephone interview or via email for major inclusion/exclusion criteria. If they were evaluated as eligible, they were invited for an in-person interview with study investigators. Participants were debriefed about the study and the effects and risks of the substance and signed informed consent. All participants underwent a physical and psychiatric examination. Volunteers with a psychiatric disorder (according to ICD-10), as well as any family history of a psychotic disorder (up to second-degree relatives) and volunteers with major physical disorders, were excluded. The major exclusion criteria included any current psychiatric disorder (according to ICD-10, excluding tobacco addiction), as well as any family history of a psychotic disorder (up to second-degree relatives), major physical disorders, pregnancy, presence of ferromagnetic materials in their body, cardio-stimulator, and left-handedness.

The whole clinical trial consisted of two arms, the EEG and fMRI arm, while each participant was expected to participate in four dosing/experimental days: two sessions with EEG (one with placebo and one with psilocybin) and two with fMRI (one with placebo and one with psilocybin). Each session involved a single dose of either psilocybin (0.26 mg/kg) or a placebo (tritici amylum) in a cross-over, double-blind manner, with a minimum of 28 days separating each session. Participants were physically re-examined and debriefed before participating in each session. The music experiments shown here were conducted in the EEG arm of the study. Participants were administered the substance in a comfortable, decorated living room–like setting, accompanied by a gender-balanced pair of sitters: at least one study clinician (psychiatrist), and a second sitter who was either a psychologist or a psychiatrist. Music was presented during the plateau phase in psilocybin sessions and during the corresponding post-administration time window in placebo sessions.

### Participants

40 healthy volunteers were enrolled in the study (20 males, 20 females) with a mean age at the start of the experiments of 35.8 (SD = 7.6). Of the entire study sample, four participants had completed high school as their highest level of education, while all remaining participants held university degrees. The first part of the experiment initially involved 20 participants who were asked to evaluate eight different compositions from various music genres to measure music-induced affective valence. However, since one participant did not complete both sessions, data from only 19 participants were analyzed. This group had a mean age of 35.9 years (SD = 7.1) at the beginning of the experiments and consisted of 10 males and 9 females, with ages ranging from 28 to 48 years. All except two participants, who had completed high school as their highest level of education, held university degrees.

From this initial evaluation, two compositions were selected based on the most pleasurable feelings and the highest contrast between placebo and psilocybin effects. For the subsequent phase of the experiment, the second half, consisting of 20 participants, listened only to these two compositions. Of the 20 participants, data from 17 were included in the final analysis: 9 were male and 8 were female, with a mean age of 36.5 years (SD = 8.3; range: 28–53 years). The remaining three participants were excluded. These exclusions were due to one participant refusing to cooperate following intense drug effects during the psilocybin session, and two others providing incomplete responses on the GEMS-9 scale, again most likely due to the overall intensity of effects. All but one participant, who had completed high school, held a university degree as their highest level of education.

### Selected compositions in the first experiment

Eight compositions were chosen to represent different genres and subgenres of music, among them metal/industrial metal and rock/punk, which are loud and intense styles characterized by distorted electric guitars, powerful drumming, and often aggressive vocals that may push emotional boundaries. The selection also included cross-cultural folk/relaxing music, classical music commonly used in psilocybin-assisted therapy sessions and controlled settings, and electronic dance subgenres associated with modern psychedelic use at raves and parties. Genre and subgenre labels were assigned using the community-sourced databases Discogs [33] and MusicBrainz [34] reflecting listener-based categorization. For a detailed description of the selected music compositions, please refer to Table 1. The first two minutes of each composition were presented in order to prevent bias from the different durations of each composition.

**Table 1.** Eight music compositions chosen to pilot affective valence to music in the first experiment, with their respective genres/subgenres and artists.

| Music genre/subgenres | Artist | Composition |
| --- | --- | --- |
| Rock/Punk | Sex Pistols | Anarchy in the UK |
| Metal/Industrial Metal | Ministry | Thieves |
| Classical/Baroque | William Boyce | Symphony No. 5 in D Major:<br>II. Tempo di Gavotta |
| Electronic/Dub | Dreadzone | Cave of the Angels |
| Folk/Nordic Folk | Hedningarna | Juopolle Joutunut |
| Electronic/Techno | Underworld | Born Slippy |
| Electronic/Trance | Delerium Feat. Sarah McLachlan | Silence |
| Electronic/Psytrance | Parasense | Gidra |

Note. Spotify links for each composition: Anarchy in the UK, Thieves, Symphony No. 5 in D Major: II. Tempo di Gavotta, Cave of the Angels, Juopolle Joutunut, Born Slippy, Silence, Gidra

### Selected compositions in the second experiment

Based on the first experiment, two compositions representing Classical and Psytrance were selected for detailed analysis. The Classical composition was selected as the composition with the highest overall affective valence, reflecting general aesthetic appeal, whereas the Psytrance composition was selected as the composition most responsive to psilocybin. Classical music is characterized by its structured forms and established conventions, typically including chamber music, solo performances, and orchestral compositions [35]. Psytrance is noted for its synthesized sounds, high tempos, repetitive beats, and layered melodies [36,37]. Although Psytrance is technically a subgenre of Electronic music, we use the simplified label Psytrance in the main analysis to clearly indicate the specific style tested. Participants listened to the two compositions in 1) full length in the case of Psytrance, approximately 8 minutes, and 2) in the case of Classical, we have looped the composition twice so it has roughly the same duration as Psytrance. The composition “Symphony No. 5 in D Major: II. Tempo di Gavotta” is in C Major and plays at 123 BPM. It exhibits low energy (8), moderate happiness/valence (61), and high instrumentalness (77), and is notably acoustic (acousticness: 94). “Gidra” by Parasense is in G Major and plays at 146 BPM. It features high energy (82), moderate happiness/valence (58), and very high instrumentalness (89), with no acoustic elements (acousticness: 0). These values are reported by Tunebat, which presents audio features derived from Spotify’s Web API on a normalized scale from 0 to 100, where higher values indicate a stronger presence of the measured attribute [38]

## Tasks

### Likert scale for music-induced affective valence

Participants rated music-induced affective valence on a Likert-type scale ranging from −3 (strong dislike/very unpleasant) to +3 (strong like/very pleasant) after listening to each composition. The exact value as a continuous variable was then extracted using a ruler.

### Geneva Emotional Music Scale (GEMS-9)

The Geneva Emotional Music Scale (GEMS-9) was administered just after participants finished listening to each of the music compositions. This scale is based on subjective ratings ranging from 1-5 (not at all – 1, somewhat – 2, moderately – 3, quite a lot – 4, very much – 5) of nine music-induced emotions identified by [39], each representing a distinct emotional quality: wonder (amazed admiration), transcendence (spiritual elevation), power (strong triumph), tenderness (loving warmth), nostalgia (melancholic reminiscence), peacefulness (calm serenity), joyful activation (energetic happiness), sadness (sorrowful longing), and tension (nervous agitation). We also computed a GEMS total score for each participant by summing their ratings across the nine emotion categories, providing an index of overall emotional responsiveness to music.

### Data analysis

In the first experiment, 19 participants were analysed, each contributing repeated ratings across 2 treatment conditions and 8 musical compositions. Descriptive analyses were used to identify (i) the composition rated as most liked overall across psilocybin and placebo conditions and (ii) the composition showing the largest psilocybin–placebo difference in ratings. A repeated-measures ANOVA was used to test whether the effect of treatment condition on music-induced affective valence differed across compositions (Condition × Composition interaction). The model included treatment condition (psilocybin vs placebo) and musical composition (eight compositions), with sequence (psilocybin-first vs placebo-first) and period (session number) included to account for potential order effects. Where a significant Condition × Composition interaction was observed, exploratory Newman– Keuls post hoc comparisons were used to characterize the interaction pattern. The significance threshold was set at α = 0.05.

In the second experiment, scores on the GEMS-9 subscales, GEMS-9 total score, and music-induced affective valence scores were evaluated using a linear mixed model (LMM) to account for individual differences where equal variances across conditions cannot be assumed. Analyses included n = 17 participants, each contributing data under 2 drug conditions and 2 musical compositions. Fixed main effects of Composition (Classical, Psytrance), Condition (Psilocybin, Placebo), and their interaction were evaluated. Random effects of individual changes between the psilocybin placebo condition were accounted for. We controlled for a randomization order (placebo/psilocybin first), age, and dose of psilocybin. Models including age and dose of psilocybin did not explain significantly more variance and were dropped from the final model. Statistical analysis was performed using R in R Studio. LMM was employed using the R base function aov() wrapper of the linear model. The random effects term was included: Score ∼ Composition × Condition + Error (ID / (Composition × Condition) + Order, where “Condition” refers to the drug manipulation (psilocybin vs. placebo), and the error term accounts for repeated measures nested within participants and music compositions. The significance threshold was set at α = 0.05. Separate models were fitted for each of the nine GEMS-9 subscales, the GEMS-9 total score, and music-induced affective valence.

P-value was obtained by comparing the linear mixed model empirical (observed) statistics for the main effects and interaction to a distribution of permutation-based statistics (obtained from 10,000 iterations using the sample() function) and computed as a percentile of permutations with lesser values. Permutation testing offers a robust non-parametric alternative to parametric inference. Unlike ANOVA, which relies on assumptions about the underlying distribution of the data (e.g., normality and homogeneity of variance), permutation methods construct a null distribution empirically by resampling the observed data. This approach therefore avoids violations of distributional assumptions and provides more accurate control of the familywise Type I error rate [40,41]. Permutation testing controlled the Type-I error rate for the fixed effects (Composition, Condition, and their interaction) within each model. Given the exploratory, hypothesis-generating nature of the study, no correction for multiple comparisons across GEMS-9 subscales was applied.

## Results

### Music-induced affective valence (Likert scale)

Across both drug conditions, the Classical/Baroque composition showed the highest mean affective valence of M = 1.94, SE = 0.19. A repeated-measures ANOVA revealed a strong main effect of musical composition on music-induced affective valence, F(7,119) = 6.637, p < 0.001, no significant main effect of treatment condition F(1,17) = 3.623, p = 0.074, and a significant Condition × Composition interaction, F(7,119) = 2.225, p = 0.037, indicating that the effect of psilocybin depended on the musical composition. No significant effects of sequence (order of conditions) or period (session number) were observed.

Descriptive analyses indicated that psytrance exhibited the largest average psilocybin–placebo difference (Δ = +0.79). Exploratory Newman–Keuls post hoc comparisons suggested that Electronic/Psytrance (p = 0.028) and Metal/Industrial Metal (p = 0.023) showed greater affective valence under psilocybin relative to placebo. For more detailed information, see Fig. 1a and Fig. 1b.

**Fig. 1.**
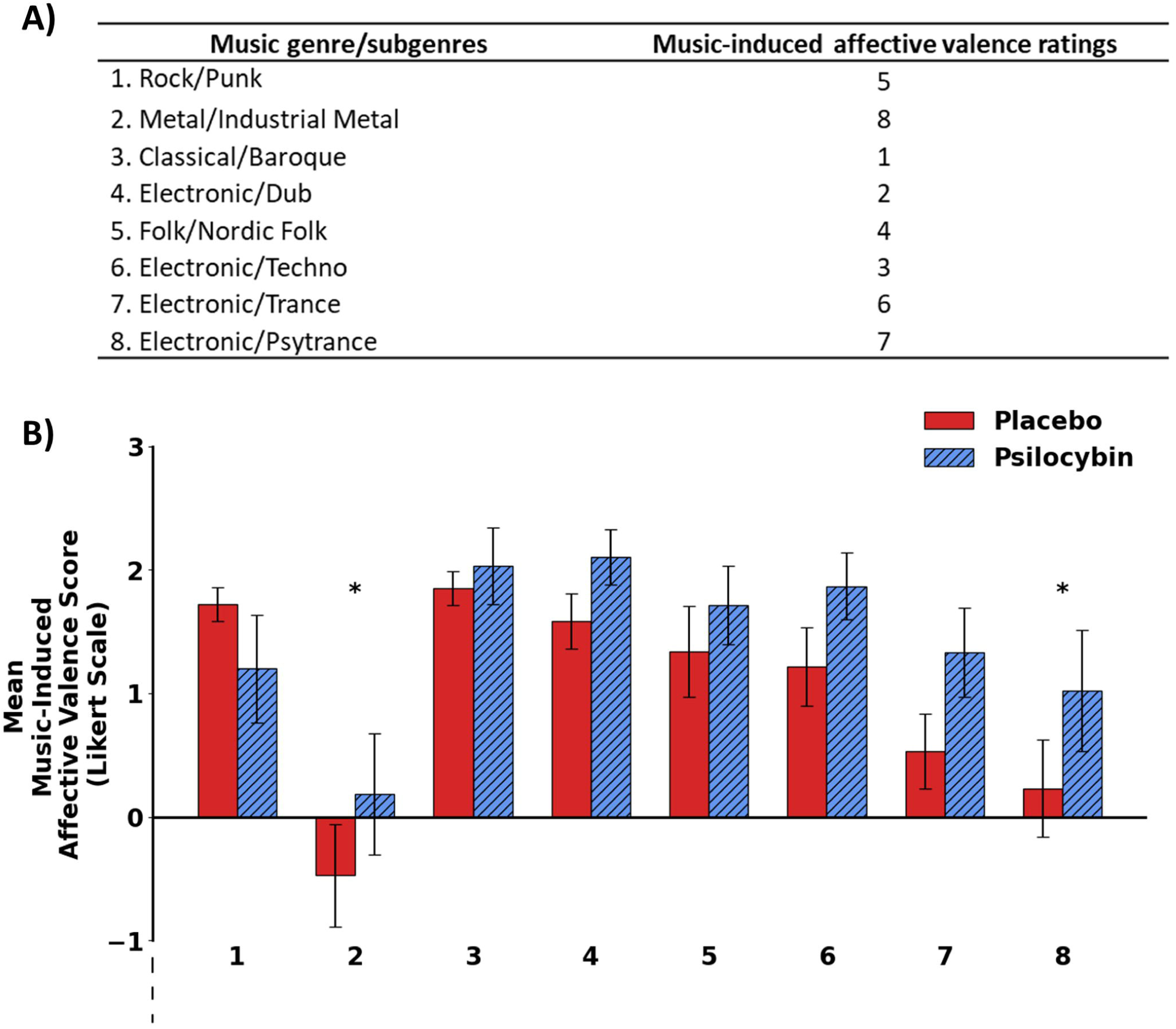
**(a)** Music-induced affective valence ratings for eight compositions from different genres in the first experiment. **(b)** Mean music-induced pleasure scores for the same compositions under psilocybin and placebo conditions. Numbers in Fig. 1b correspond to the genre/subgenre labels shown in the left column of Table 1a. (*p < 0.05; error bars represent SEM. Faded dashed lines indicate that the scale extends to –3.)

In the second experiment (2 compositions), there were no significant effects of composition or the interaction between compositions and conditions on music-induced affective valence. However, there was an effect of the condition. Despite both groups indicating relatively low levels of music-induced affective valence, psilocybin significantly increased the affective valence derived from psilocybin (M = 1.76, SE = 0.26 *vs*. 1.05, SE = 0.20, p = 0.017) compared to the placebo condition. For more detailed information, see Figs. 2a-c

**Fig. 2.**
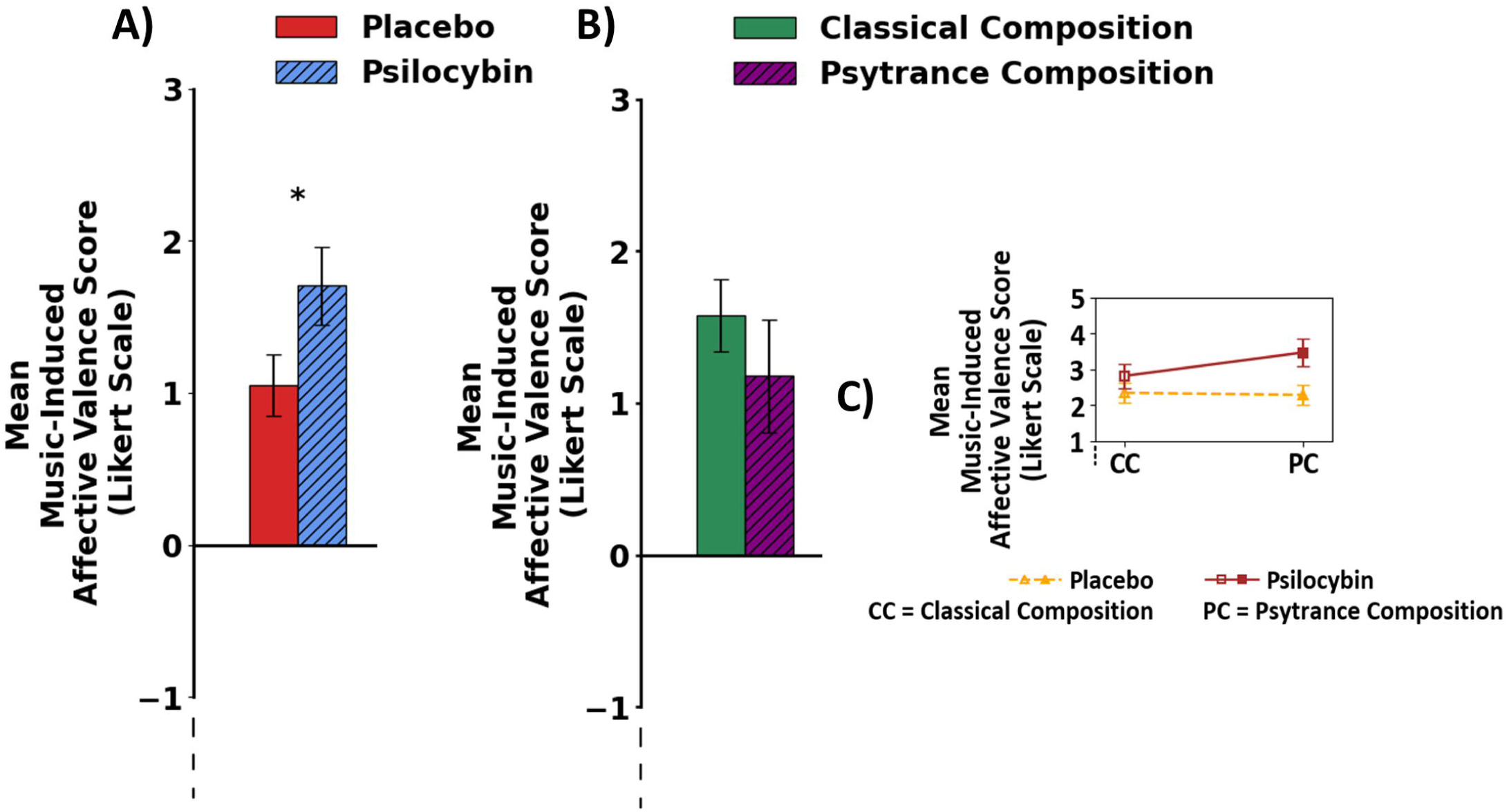
**(a)** Mean music-induced affective valence scores in the second experiment under psilocybin and placebo conditions (collapsed across compositions). **(b)** Mean music-induced affective valence scores for classical vs. psytrance compositions (collapsed across conditions). **(c)** Interaction effect of composition and condition on mean music-induced affective valence scores. *p < 0.05; error bars represent SEM. Faded dashed lines indicate that the scale extends to −3

### Effects of treatment condition (psilocybin vs. placebo) on GEMS-9

The analysis revealed a significant effect of condition on three factors from the GEMS-9, with psilocybin significantly increasing wonder, transcendence, and joyful activation compared to the placebo. Specifically, participants in the psilocybin condition reported statistically higher wonder mean score than those in the placebo group (M = 3.15, SE = 0.31 vs. M = 2.32, SE = 0.17; p = 0.012). Similarly, the transcendence scores were significantly higher in the psilocybin group (M = 2.71, SE = 0.24) compared to the placebo group (M = 1.91, SE = 0.18), showing a significant effect (p = 0.011). For joyful activation, the psilocybin group also demonstrated significantly (p = 0.015) higher mean score (M = 3.15, SE = 0.24) than the placebo condition (M = 2.50, SE = 0.16). No significant differences were observed for the remaining GEMS-9 factors. For more detailed information, see Fig. 3a.

**Fig. 3.**
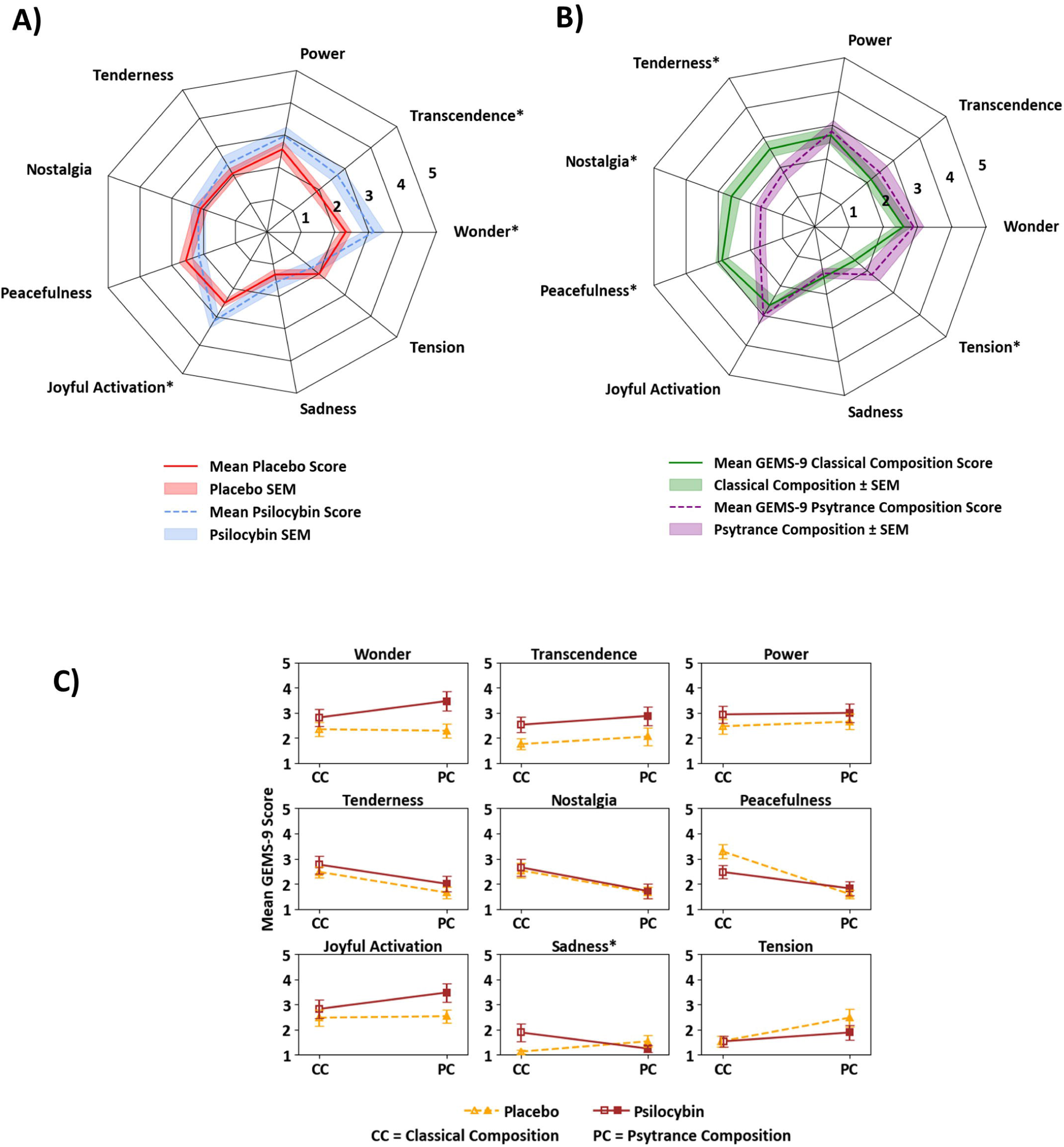
**(a)** Mean GEMS-9 subscale scores for psilocybin vs. placebo (collapsed across classical and psytrance compositions). **(b)** Mean GEMS-9 subscale scores for classical vs. psytrance compositions (collapsed across psilocybin and placebo conditions). **(c)** Interaction effects of composition (classical vs. psytrance) and condition (psilocybin vs. placebo) on mean GEMS-9 subscale scores. * indicates statistical significance (P < 0.05)

### Effects of composition (classical vs. psytrance) on GEMS-9

Out of the nine emotions measured by GEMS-9, a significant effect of composition (classical vs. psytrance) was found in nostalgia, tenderness, peacefulness, and tension. The classical composition evoked significantly greater nostalgia compared to the psytrance composition, as shown by higher mean scores (M = 2.59, SE = 0.26) versus (M = 1.68, SE = 0.16), (p = 0.008). Similarly, the classical composition increased tenderness (M = 2.62, SE = 0.24) relative to the psytrance composition (M = 1.82, SE = 0.23) (p = 0.028). Peacefulness was also more pronounced in response to the classical composition (M = 2.88, SE = 0.19) compared to the psytrance composition (M = 1.71, SE = 0.18), yielding the most significant effect (p < 0.001). In contrast, tension was significantly greater under the psytrance composition (M = 2.18, SE = 0.28) compared to the classical composition (M = 1.53, SE = 0.17) (p 0.05). No significant differences were observed for the remaining GEMS-9 factors. For more detailed information, see Fig. 3b.

### Interaction between compositions and condition (GEMS-9)

A significant interaction effect was observed between condition and composition on music-evoked sadness, p = 0.039. Specifically, under the placebo condition, sadness levels were higher when participants listened to the psytrance composition (M = 1.53, SE = 0.23) compared to the classical composition (M = 1.12, SE = 0.08). However, under psilocybin, the pattern reversed, and sadness was higher when listening to the classical composition (M = 1.88, SE = 0.35) compared to the psytrance composition (M = 1.24, SE = 0.14). No significant condition-by-composition interactions were observed for the other GEMS-9 factors. For more detailed information, see Fig. 3c.

### GEMS-9 Total Score

There were no significant effects of either music composition or condition, nor was there an interaction between composition and condition on the GEMS total score.

## Discussion

In the first experiment, the classical composition showed the highest mean affective valence across both treatment conditions, while the psytrance composition exhibited the largest psilocybin–placebo difference. This effect was confirmed in the second experiment. We found no main effects of composition, condition or their interaction on the GEMS-9 total score. Psilocybin did, however, selectively enhance specific music-induced emotions, wonder, transcendence, and joyful activation, across both classical and psytrance compositions. Composition itself shaped the profile: relative to each other, the classical composition elicited higher tenderness, peacefulness, and nostalgia, whereas psytrance evoked more music-induced tension in both psilocybin and placebo conditions. There was a composition × drug interaction for sadness: under placebo, sadness was higher with psytrance than classical, but under psilocybin, this pattern reversed, with classical eliciting more sadness than psytrance.

### Music-induced affective valence

Although not all effects reached statistical significance, psilocybin enhanced music-induced affective valence for most compositions representing different genres across both experiments, with the exception of the rock/punk composition in the first experiment. The lack of enhancement for rock/punk may reflect acoustic or emotional characteristics of this genre, such as its typically short, raw, and confrontational qualities, which may be less responsive to the psilocybin-induced state [42]. It is also notable that, in the first experiment, psilocybin significantly increased affective valence for some of the least-liked compositions, particularly psytrance and industrial metal/thrash, with the latter even shifting from negative to positive ratings. This pattern suggests that psilocybin may partially elevate affective responses even to initially non-preferred music. In controlled psychedelic sessions settings, where individual musical preferences cannot always be fully accommodated, this finding may indicate that non-preferred music does not necessarily result in negative affective responses in this context.

### Emotional Responses to Music: Psilocybin vs. Placebo Condition

Psilocybin-related increases in wonder, transcendence, and joyful activation correspond to core features of mystical-type experiences as previously described [43]. In controlled settings with healthy volunteers, such psilocybin-induced mystical experiences have been linked to increased Openness [44] and lasting enhancements in personal meaning and spiritual significance, with effects reported beyond one year [45]. Given that psilocybin, when combined with music, may intensify these experiences, and considering their association with enduring positive psychological outcomes, it can be hypothesized that psilocybin, within a musical context, whether a classical or a psytrance composition is used, may serve as an amplifier of these long-term positive effects.

Our results partially overlap with findings from healthy volunteers under LSD who listened to neo-classical/ambient music [17] during the peak experience, particularly in the domains of wonder and transcendence, and extend them by showing that similar effects are observed across both classical and psytrance compositions. However, our findings differ from those in a psilocybin study involving depressed patients [18], where music-evoked peacefulness was enhanced and sadness reduced following treatment. In that study, emotional responses to music were assessed before treatment and one day after the final dosing session (10 mg followed by 25 mg), in an fMRI scanning context, whereas our study assessed music-evoked emotions during the acute psychedelic state in a placebo-controlled comparison, making the designs not directly comparable. However, these findings collectively point to a set of potential modulators of music-evoked emotions under psychedelics: population (healthy vs. clinical), timing (acute vs. post-treatment), dosage regimen, setting (scanner vs. therapeutic context), and, to a lesser extent, the specific psychedelic compound and whether measurement occurs at peak or plateau.

### Emotional Responses to Music: Classical vs. Psytrance Composition

The observed increase in peacefulness, tenderness, and nostalgia for the classical composition, all of which are low-arousal, high-valence emotions [46], is consistent with the fact that these states are commonly elicited by classical music, have previously been reported more frequently in response to classical music than to electronic genres such as techno, and are understood as introspective states [39]. Music-induced nostalgia in particular serves important psychological functions as it fosters social connectedness, enhances self-esteem, youthfulness, optimism, and inspiration, and strengthens meaning in life and self-continuity [47]. In contrast, psytrance elicited music induced tension, a high arousal, low valence emotion [46], which can arise from uncertainty and unmet expectations [48] such as those created by the layered melodies characteristic of the psytrance genre [36,37]. Overall, this pattern shows that, relative to each other, the two compositions evoke genre-appropriate and distinct emotional profiles, consistent with evidence that low-arousal, high-valence and high-arousal, low-valence emotions are associated with largely distinct neural activation patterns within the GEMS-9 framework [46]. The classical composition was associated with affective states that may be more readily aligned with those typically targeted in therapeutic or supported psychedelic sessions, and this pattern was observed under both placebo and psilocybin conditions. This represents a potentially relevant observation, suggesting that control/placebo sessions in this type of paradigm may not necessarily be affectively neutral, and highlighting musical composition as a potentially important determinant of the affective experience.

### Interaction between musical compositions and conditions

Although overall sadness ratings were low, classical composition under the influence of psilocybin elicited higher levels of music-induced sadness compared to the psytrance composition. Sadness, previously identified as a component of challenging psychedelic experiences, has also been associated with beneficial psychological processes such as self-reflection [49] and emotional breakthrough [50], particularly in observational studies involving psychedelics, including psilocybin. This might be especially relevant given that music-evoked sadness has been described as more melancholic than overtly sad or depressed, which may reflect a more contemplative and less distressing emotional tone than that of everyday sadness [39]. A recent analysis of the long-term effects of psilocybin in volunteers from this study found that individuals who experienced moments of emotional challenge, such as anxiety or discomfort within otherwise mixed emotional experiences in the EEG arm, reported more favorable long-term subjective outcomes than those with exclusively positive sessions [32]. Taken together, if psilocybin-facilitated music experiences that evoke emotional challenges, such as sadness, can contribute to constructive emotional processes or beneficial long-term outcomes, as suggested by previous research, it is plausible to consider, based on our findings, that classical music may support these psychological benefits more than psytrance during the plateau phase of psilocybin’s effects.

Our data also suggest that, under psilocybin, psytrance was associated with lower sadness ratings compared to classical music during the plateau phase. It would be interesting to examine whether similar effects occur during other phases of the psychedelic experience, particularly the come-up, which is often described as stressful and characterized by negative emotional states [51]. Given that music-evoked sadness has been linked to challenging psychedelic experiences [49], if a similar pattern were observed during this stage, reduced sadness could potentially support participants in navigating the early phase.

Apart from sadness, none of the other interaction effects reached statistical significance, and overall emotional differences across conditions were relatively subtle. Interestingly, although the two compositions differed in genre, both were characterized by higher tempo, major key, moderate happiness, and high instrumentalness, which may have contributed to the relatively subtle interaction effects observed. Notably, they differed more clearly in acoustic properties and energy. Future studies may benefit from directly comparing musical features such as tempo, tonality, vocal presence, or emotional valence within genres to better understand their role within the psychedelic context. Additionally, while a controlled setting is more typical for classical music in psychedelic sessions [28], psytrance is usually experienced in a dynamic, naturalistic environment with an emphasis on the communal aspect [52] and therefore may not have fully expressed its impact within the individually focused, structured context of the experiment. It is also worth noting that we presented short musical pieces, approximately 8 minutes in length, and that the overall effect of the session is likely influenced by the longer duration of the session itself and the cumulative impact of the music.

### Hypothetical underlying neurobiological mechanisms

Music-induced chills, brief peak arousal episodes, are associated with striatal activation [53] and dopamine release in the ventral striatum [54]. Psilocybin has been shown to acutely increase dopamine release in the dorsal striatum, with a ventral trend linked to euphoria [55], suggesting a dopaminergic pathway through which it could amplify music-induced affective valence for compositions already rated positively. In parallel, a recent fMRI study found that psilocybin modulates effective connectivity within the fronto–striatal–thalamic circuit [56], strengthening bottom-up signals from the thalamus and striatum and reducing top-down influence from the cortex to these subcortical regions. The striatum is central to reward processing [57], the thalamus serves as a sensory gateway [58], and parts of the frontal cortex, particularly the orbitofrontal and ventromedial prefrontal cortices, have been shown to encode subjective preferences by representing the expected value and experienced pleasure of stimuli [59]. In that study, bottom-up connectivity increases were strongest in subcortical regions with high D receptor density, consistent with preferential engagement of dopaminergic pathways. If similar connectivity shifts occur acutely alongside psilocybin-induced striatal dopamine release, they may temporarily elevate the reward value of musical input, even for compositions typically rated low in music-induced affective valence, by making the circuit more responsive to reward signals and less constrained by established preferences. At the same time, the absence of a psilocybin-related boost in introspective music-evoked emotions (peacefulness, tenderness, nostalgia) across compositions is consistent with evidence that psilocybin reduces DMN integrity and mPFC– PCC coupling, a network implicated in self-referential mentation [60–62].

### Limitations and recommendations

Although this study provides valuable preliminary insights, as it was exploratory in nature and embedded within a larger clinical trial, several limitations should be considered when interpreting the findings and designing future research. Familiarity with the musical compositions can enhance subjective valence ratings [63] and evoke autobiographical memories that intensify emotional responses [64,65], while musical preferences, defined as genre or song liking, have also been shown to shape emotional responses to music [39,63]. We did not assess the genre liking and familiarity, and although we measured music-induced affective valence and emotional responses under psilocybin, we did not examine their relationship, as this was beyond the study’s scope. As this is a pilot study with a small sample size, some effects may not have been detected but could become apparent with larger samples. Future research could benefit from integrating these factors in a single model with larger samples and a priori defined hypotheses to test potential mediation or moderation effects and better understand their interactions under psychedelics. Also because each musical genre was represented by a single composition, conclusions about emotional responses to music cannot be generalized to broader genre categories. Future studies should include multiple exemplars per genre to disentangle genre-level effects from composition-specific effects.

## Conclusion

To conclude, in healthy individuals within a controlled setting, psilocybin’s modulation of music-induced affective valence ranges from composition-selective to broadly amplified across different experimental conditions. Both psilocybin (across both compositions) and the compositions (representing different genres) emerged as determinants of music-evoked emotions during the plateau phase, each selectively enhancing different emotional components. Their combined effects showed limited interaction overall; however, under psilocybin, sadness increased during the classical composition but decreased during the psytrance composition.

## Data Availability

The minimal dataset underlying the results of this study will be deposited in a public repository upon acceptance for publication and will be made publicly available no later than the time of publication.

